# Systematic review protocol: Quantitative susceptibility mapping of brain iron accumulation in neurodegenerative diseases

**DOI:** 10.1101/2020.02.18.20022608

**Authors:** Parsa Ravanfar, Samantha Loi, Tamsyn Van Rheenen, Ashley Bush, Patricia Desmond, Vanessa Cropley, Bradford Moffat, Dennis Velakoulis, Christos Pantelis

**Affiliations:** Melbourne Neuropsychiatry Centre, Department of Psychiatry, The University of Melbourne and Melbourne Health, Victoria, 3053, Australia; Neuropsychiatry Unit, Melbourne Neuropsychiatry Centre, The University of Melbourne, NorthWestern Mental Health, Melbourne Health, Parkville, Victoria, Australia; Melbourne Dementia Research Centre, Florey Institute of Neuroscience and Mental Health, The University of Melbourne, Victoria, Australia; Department of Radiology, The University of Melbourne, Royal Melbourne Hospital, Parkville, VIC, Australia; Melbourne Brain Centre Imaging Unit, Department of Medicine and Radiology, The University of Melbourne, Parkville, Australia; Florey Institute of Neuroscience and Mental Health, The University of Melbourne, Victoria, Australia; Centre for Mental Health, Swinburne University of Technology, Victoria Australia

**Keywords:** quantitative susceptibility mapping, QSM, neurodegenerative diseases, iron, MRI, magnetic resonance imaging, Alzheimer disease, Parkinson disease, Huntington disease, Wilson disease, amyotrophic lateral sclerosis, dementia with Lewy bodies, vascular dementia, systematic review

## Abstract

Iron has been found to play an important role in neurodegeneration. Quantitative susceptibility mapping (QSM) is a relatively new – and the most accurate - MRI technique available for assessment of iron deposition in the brain. There is a rapidly growing number of studies using QSM to investigate brain iron distribution in neurodegenerative diseases including but not limited to Alzheimer’s disease, Parkinson’s disease, Amyotrophic lateral sclerosis, Huntington disease and Wilson’s disease. These studies have shown increased iron deposition in the brain regions that are associated with the pathology of the disease. Additionally, QSM is found to be accurate in differential diagnosis of neurodegenerative diseases where clinical presentations are indistinguishable. Structural changes evidenced by QSM are reported to precede the onset of clinical manifestation of neurodegenerative diseases suggesting its benefit in early diagnosis. To our knowledge, no systematic review of QSM findings in neurodegenerative diseases has been published before. A systematic synthesis and conclusion of the existing evidence can improve our understanding of the pathophysiology of neurodegeneration, describe the clinical and research utility of QSM, and point out the direction for future studies in neuropsychiatric disorders.

This document is a systematic review protocol developed in accordance with Preferred reporting items for systematic review and meta-analysis protocols (PRISMA-P) guideline. This protocol is prepared as a guide for conducting a systematic review of studies investigating brain iron and microstructural changes in neurodegenerative diseases using quantitative susceptibility mapping (QSM). This protocol has also been submitted to Prospective Register of Systematic Reviews (PROSPERO) for registration. By publishing this protocol, we aim to enhance clarity and transparency of our systematic review and minimise the risk of bias in the process of its development.

## Section 1, Administrative information

### Item 1. Title

A systematic review of quantitative susceptibility mapping of brain iron accumulation in neurodegenerative diseases.

### Item 1a. Identification

This report will be produced as a systematic review

### Item 1b. Update

N/A

### Item 2. Registration

This review has been submitted to be registered at PROSPERO, reference ID: 168598

### Item 3b. Contributions

PR will develop search strategies, run the database search, and in parallel with SL, each will independently perform the screening of studies for inclusion, data extraction and quality assessment. CP will adjudicate the conflicts between two reviewers, PR and SL, in screening, data extraction, and quality assessment if they arise.

Development of protocol is done by PR, and all authors will collaborate to improve and approve the final protocol.

Data synthesis will be performed by PR and SL. CP, DV, AB and BM will provide intellectual input to data synthesis and interpretation.

VC, PD, AB, BM, DV, CP, TVR and SL will provide intellectual input into the manuscript draft prepared by PR.

All authors will read, provide feedback and approve the final manuscript.

### Item 4. Amendments

Amendments to this protocol that are made after registration will be recorded and reported in the final publication.

### Item 5. Support

#### Item 5a. Sources

No funding or sources of support were received for the purpose of production of this review. All authors are either student or academic staff at The University of Melbourne.

#### Item 5b. Sponsor

N/A

#### Item 5c. Role of sponsor or funder

N/A

## Section 2. Introduction

### Item 6. Rationale

Iron plays an important role in the development and progression of neurodegenerative diseases. In the central nervous system (CNS), iron plays a crucial physiologic role in myelin synthesis and neurotransmitter production. However, excess amounts of iron can induce cytotoxic pathways in the neural cells. Intracellular accumulation of iron facilitates Fenton reaction which produces a hydroxyl radical (OH•) from break-down of hydrogen peroxide (H2O2), leading to oxidative stress. Hydroxyl radicals are highly toxic to living organisms by inducing membrane lipid peroxidation and structural alterations of proteins and nucleic acids. The series of processes initiated by iron-induced oxidative damage and ultimately leading to cell death are called ferroptosis. Ferroptosis is a unique type of programmed cell death triggered by iron-induced lipid peroxidation and generation of intra-cellular lipid radicals. Additionally, iron promotes misfolding and aggregation of insoluble proteins that are the hallmark of the neurodegenerative diseases including β-amyloid, α-synuclein, and tau protein. Further, iron has been found to become structurally incorporated in tau protein and β-amyloid thereby producing greater oxidative stress.

Histopathologic investigations have shown increased iron content in various brain regions in neurodegenerative diseases such as Alzheimer’s disease and Parkinson’s disease. *In vivo* measurement of brain iron in neurodegenerative diseases has been made possible by magnetic resonance imaging (MRI) modalities sensitive to magnetic susceptibility differences across the brain such as Susceptibility Weighted Imaging (SWI) and T2* weighted imaging. Evidence from studies using these modalities suggests increased iron accumulation in certain areas of the brain in neurodegenerative diseases compared to healthy individuals. QSM is a novel susceptibility imaging modality that can quantitatively measure the distribution of iron content in the brain with superior accuracy and sensitivity than older techniques. Comparison of iron measurements performed by QSM and post-mortem histochemical analysis has reported a significant correlation between the two measurements.

In recent years, there has been an increasing number of QSM studies investigating the content and distribution of iron in the brain in neurodegenerative diseases including Alzheimer’s disease, Parkinson’s disease, Amyotrophic Lateral Sclerosis, Huntington disease, and vascular dementia. To date, however, no systematic review of the literature on findings of QSM in neurodegenerative diseases has been published. A synthesis and critical review of the distribution of iron content in neurodegenerative diseases will provide insight into the role of iron in the pathogenesis and diagnosis of these disorders and may point the way to undertaking similar studies in neuropsychiatric disorders.

### Item 7. Objectives

The primary aim of this review is to synthesize and critically review the existing evidence of disease specific iron distribution patterns observed in neurodegenerative diseases in which brain iron has been investigated using QSM. To this end, our specific objectives are:

1. To identify which neurodegenerative diseases have been studied for brain iron distribution using QSM

1.a. To compare and contrast the results of QSM studies in each studied neurodegenerative disease to identify the brain regions where there is altered iron content compared to healthy individuals.

1.b. To identify what study-level factors are associated with altered iron content in neurodegenerative diseases

## Section 3. Methods

This systematic review will be designed and produced in accordance to Preferred Reporting Items for Systematic Reviews and Meta-Analyses: The PRISMA guideline.

### Item 8. Eligibility criteria

Eligibility criteria of this review are determined according to PICO system at two stages.

In the first stage, based on the title and abstract of the records, those meeting the following criteria will be included:

1. Published in English
2. **Design:** Empirical cross sectional or longitudinal design. Case reports, review and meta-analysis studies will be excluded.
3. **Participants:** Include a human patient group with a formal diagnosis of a neurodegenerative disease regardless of age. Studies investigating pre-clinical stages of a neurodegenerative disease in genetically predisposed population or asymptomatic subjects with positive biomarkers of the neurodegenerative disease will also be included. We will use the National Institute of Health (NIH) definition for the neurodegenerative diseases and include all diseases listed under the Medical Subject Headings (MeSH) index of neurodegenerative diseases.
4. **Investigations:** QSM MRI of the brain is used in the methodology. Further details of methodology will be verified in the second stage.

In the second stage of screening, from those included in the first stage, full-text records will be deemed eligible if, in addition to the conditions in stage 1:

1. A full-text peer-reviewed original article is available. Abstract-only records presented in conferences will not be included.
2. **Investigations:** Of interest to this review are the studies investigating brain iron in neurodegenerative diseases using *QSM imaging* with MRI scanners of any field strength (1.5, 3, 4.7, 7 tesla) as the only imaging modality, or in combination with other diagnostic techniques, provided that a report of QSM measurements in isolation is given. Studies that used other iron-sensitive imaging techniques such as SWI, T2* and R2* weighted imaging without QSM will be excluded.
3. **Comparators**: We will include studies that made a comparison of magnetic susceptibility in any brain region between neurodegenerative illness and healthy controls, or among different subtypes or clinical features of any given neurodegenerative disorder. Studies that provide QSM measurements in correlation with pathologic markers of a neurodegenerative disease (e.g. β-amyloid imaging) in absence of a healthy control group will also be reviewed. Studies that focused on the QSM technical development using a population of either healthy participants or patients without reporting any implications of QSM in relevance to the neurodegenerative disease will be excluded.
4. **Outcomes**: Since QSM is not a direct measurement of iron concentration, the value for tissue magnetic susceptibility is used as a proxy of iron accumulation in the brain. Studies have used the terms “iron content/concentration” and “magnetic susceptibility” interchangeably when interpreting the findings of QSM imaging. However, the outcome measure of mean magnetic susceptibility (**χ**) in ROIs should be reported in ppm (parts per million) or ppb (parts per billion).
5. Presented results are from an independent sample of data. If there are more than one studies that have used the same sample (re-analysed the same dataset or sample population), the first publication will be included in the systematic review if eligible according to other criteria.

### Item 9. Information sources

PubMed, Embase, Scopus, and PsycInfo databases will be searched to access all relevant literature published as of Feb 2020. Relevant review studies will also be checked for their references to ensure an exhaustive search has been performed and to minimize the risk of missing any studies. If required, authors of included studies will be contacted to obtain additional information.

### Item 10. Search strategy

For each individual database interface, a relevant search strategy is developed to account for technical variations. Considering the focus of this review being QSM findings in neurodegenerative diseases, we will first perform a search containing QSM or “quantitative susceptibility mapping” and MeSH heading of “neurodegenerative diseases” in PubMed which maps the search to include all subheadings under the term “neurodegenerative diseases”. This search will include all results that have been indexed under any of the subheadings of neurodegenerative diseases.

In order to include all studies that contain the name of any of the neurodegenerative diseases in their text, in the next step, all MeSH subheadings under “neurodegenerative disease” will be copied in a new search query as a free-text search separated by “OR”. This search query will then be combined with QSM or “quantitative susceptibility mapping” query. The same process will be repeated with each neurodegenerative disease name modified to contain all varieties of the term. For example, for the MeSH term “Alzheimer disease”, in the modified search, we will use Alzheimer* to include Alzheimer’s disease, Alzheimer, and Alzheimer dementia.

In the third step, in all databases, we will search for neurodegen* AND (QSM OR “quantitative susceptibility mapping”) to ensure we have gathered all studies that contain terms related to neurodegeneration and neurodegenerative.

The specific final search queries for all databases will be published as an appendix to this review article.

This search will be repeated once again at the end of data extraction phase with a date limit between the initial search and the new date to ensure inclusion of the new published studies.

### Item 11. Study records

#### Item 11a. Data management

The outcome of search strategies described above, will be imported in Mendeley reference manager software, Elsevier, London, UK. Full citations including title, authors, abstract, keywords, journal name and pages, as well as digital identifiers like DOI and URL will be stored. References will be checked for duplicates, to merge the duplicate studies. The resultant reference list will be imported into Covidence online tool, Melbourne, Australia for further inclusion assessments.

Full text articles of all studies that pass the title and abstract screening according to the eligibility criteria will be obtained and added to the records in Mendeley.

#### Item 11b. Selection process

Selection of studies for inclusion will be performed in two stages. In the first stage, titles, abstracts, and keywords will be examined to exclude irrelevant studies. For studies without an online abstract, a full text article will be used. As the result of the first stage, studies will be categorized as eligible, not eligible, and potentially eligible. Potentially eligible studies are those records in which a clear decision of their inclusion cannot be made solely by assessment of their title and abstract.

In the second stage, full-text articles of eligible or potentially eligible studies will be carefully reviewed and selected according to their fulfillment of the review criteria. The reasons for exclusion of each study at this stage will be recorded and reported.

PR and SL will independently perform these two stages of selection process for the studies imported in Covidence. For each study to be included, it is required to be voted in favour of inclusion by both reviewers. In instances of a conflict between the two reviewers’ decisions, a consensus will be reached by a discussion between reviewers. If the disagreement persists, a third author (DV) will assist to determine the inclusion or exclusion of that study.

#### Item 11c. Data collection process

Extraction of data from included studies will be independently performed by two reviewers in parallel (PR and SL). Data extraction forms that are collaboratively made by the authors to extract data from studies will be used. This form will be published as an appendix to this review. Discrepancies between data extraction performed by two authors will be discussed to reach an agreement. Unresolved disagreements will be adjudicated by a third author, DV.

### Item 12. Data items

Data items that are relevant to the purpose of this review include:

- Neurodegenerative disease investigated
- Sample characteristics: Age, Sex, sample size, disease duration, and disease subtype where relevant
- Investigation characteristics: MRI scanner field strength, modalities used other than QSM, regions of interest (ROIs) investigated
- Outcomes: increased, decreased, or unchanged susceptibility (iron content) in each ROI, and its correlation with age, disease duration, and clinical features, as well as correlation of QSM with other iron measurements such as histochemical iron analysis, and other susceptibility sensitive modalities. Statistical effect size and p-value will be reported.

### Item 13. Outcomes and prioritization

Primary outcomes:

To identify disease-specific alterations of brain iron distribution in various neurodegenerative diseases. To this aim, specific outcomes of interest will be:

- Identification of the neurodegenerative diseases that have been studied for brain iron distribution using QSM.
- In each studied neurodegenerative disease, identification of the regions of the brain that show altered iron content.
- Identification of whether alteration of brain iron content correlates with pathological processes (e.g. amyloid or tau related pathology) and the brain regions involved in each given disease.
- Identification of whether alteration of brain iron content correlates with clinical features of the disease such as duration, severity, and subtypes. This will provide insight into whether iron dyshomeostasis in the brain is the cause, consequence, or an independent pathway for neurodegeneration.

### Item 14. Risk of bias in individual studies

A quality assessment for risk of bias will be performed on each record included in this review. We will use the National Heart, Lung and Blood Institute (NHLBI) Tool for Assessment of the Quality of Case-Control Studies for quality assessment of reviewed studies. In order to improve the application of this tool to the studies focused in this review, minor adjustments will be made in the items of the questionnaire. We will publish the utilized checklist as an appendix to the final manuscript.

In order to minimize the risk of bias in data extraction process, quality assessment of studies will be performed after data extraction is completed.

### Item 15. Data synthesis

In a narrative synthesis, we will provide a consolidated report and interpretation of alterations in the magnetic susceptibility and iron content in various brain regions of each neurodegenerative disease. We aim to provide a critical discussion based on the results of reviewed studies to answer the objectives outlined previously.

For a quantitative synthesis of the results, mean susceptibility values of brain regions investigated in each neurodegenerative disease will be the main numerical outcome measure. According to our scoping search, due to differences in measurement of susceptibility, MRI scanner field strength and variation of reference regions considered for QSM in different studies, it is unlikely that a quantitative analysis of the reported susceptibility values can be performed. However, a quantitative synthesis or meta-analysis will be pursued if it is deemed possible after extraction of data.

### Item 16. Meta-bias(es)

In order to minimize the “outcome reporting” bias, we will meticulously check and record the ROIs investigated in the methodology of included studies and compare them with the reported results. By this approach, we will be able to take into account all the instances that the null hypothesis could not be rejected. If possible, Funnel plots will also be used to investigate the potential for publication/dissemination bias.

### Item 17. Confidence in cumulative evidence

In this review, our confidence in the accumulative evidence will be determined and reported according to the GRADE (Grading of Recommendations, Assessment, Development and Evaluations) approach. Where applicable, the quality of evidence will be evaluated within the domains of risk of bias, imprecision, inconsistency, indirectedness, and publication bias.

## Data Availability

The manuscript is a protocol for a systematic review. No empirical data is presented in this manuscript.

## Appendix 1. Search strategy

PubMed:

1. (QSM OR “quantitative susceptibility mapping”) AND (neurodegenerative diseases[MeSH Terms])
2. Disease names as appear under MeSH heading for “neurodegenerative diseases”: (QSM OR “quantitative susceptibility mapping”) AND ((“Chronic Traumatic Encephalopathy”[All Fields] OR “Heredodegenerative Disorders, Nervous System”[All Fields] OR “Alexander Disease”[All Fields] OR “Amyloid Neuropathies, Familial”[All Fields] OR “Bulbo-Spinal Atrophy, X-Linked”[All Fields] OR “Canavan Disease”[All Fields] OR “Cockayne Syndrome”[All Fields] OR “Dystonia Musculorum Deformans”[All Fields] OR “Gerstmann-Straussler-Scheinker Disease”[All Fields] OR “Hepatolenticular Degeneration”[All Fields] OR “Hereditary Central Nervous System Demyelinating Diseases”[All Fields] OR “Hereditary Sensory and Autonomic Neuropathies”[All Fields] OR “Dysautonomia, Familial”[All Fields] OR “Hereditary Sensory and Motor Neuropathy”[All Fields] OR “Alstrom Syndrome”[All Fields] OR “Charcot-Marie-Tooth Disease”[All Fields] OR “Giant Axonal Neuropathy”[All Fields] OR “Refsum Disease”[All Fields] OR “Spastic Paraplegia, Hereditary”[All Fields] OR “Huntington Disease”[All Fields] OR “Lafora Disease”[All Fields] OR “Myotonia Congenita”[All Fields] OR “Myotonic Dystrophy”[All Fields] OR “Neurofibromatoses”[All Fields] OR “Neurofibromatosis 1”[All Fields] OR “Neurofibromatosis 2”[All Fields] OR “Neuronal Ceroid-Lipofuscinoses”[All Fields] OR “Optic Atrophies, Hereditary”[All Fields] OR “Optic Atrophy, Autosomal Dominant”[All Fields] OR “Optic Atrophy, Hereditary, Leber”[All Fields] OR “Wolfram Syndrome”[All Fields] OR “Pantothenate Kinase-Associated Neurodegeneration”[All Fields] OR “Spinal Muscular Atrophies of Childhood”[All Fields] OR “Spinocerebellar Degenerations”[All Fields] OR “Friedreich Ataxia”[All Fields] OR “Myoclonic Cerebellar Dyssynergia”[All Fields] OR “Olivopontocerebellar Atrophies”[All Fields] OR “Spinocerebellar Ataxias”[All Fields] OR “Machado-Joseph Disease”[All Fields] OR “Tourette Syndrome”[All Fields] OR “Tuberous Sclerosis”[All Fields] OR “Unverricht-Lundborg Syndrome”[All Fields] OR “Motor Neuron Disease”[All Fields] OR “Amyotrophic Lateral Sclerosis”[All Fields] OR “Bulbar Palsy, Progressive”[All Fields] OR “Muscular Atrophy, Spinal”[All Fields] OR “Bulbo-Spinal Atrophy, X-Linked”[All Fields] OR “Spinal Muscular Atrophies of Childhood”[All Fields] OR “Paraneoplastic Syndromes, Nervous System”[All Fields] OR “Anti-N-Methyl-D-Aspartate Receptor Encephalitis”[All Fields] OR “Limbic Encephalitis”[All Fields] OR “Myasthenia Gravis”[All Fields] OR “Lambert-Eaton Myasthenic Syndrome”[All Fields] OR “Myelitis, Transverse”[All Fields] OR “Opsoclonus-Myoclonus Syndrome”[All Fields] OR “Paraneoplastic Cerebellar Degeneration”[All Fields] OR “Paraneoplastic Polyneuropathy”[All Fields] OR “Postpoliomyelitis Syndrome”[All Fields] OR “Prion Diseases”[All Fields] OR “Encephalopathy, Bovine Spongiform”[All Fields] OR “Gerstmann-Straussler-Scheinker Disease”[All Fields] OR “Insomnia, Fatal Familial”[All Fields] OR “Kuru”[All Fields] OR “Scrapie”[All Fields] OR “Wasting Disease, Chronic”[All Fields] OR “Subacute Combined Degeneration”[All Fields] OR “Synucleinopathies”[All Fields] OR “Lewy Body Disease”[All Fields] OR “Multiple System Atrophy”[All Fields] OR “Parkinson Disease”[All Fields] OR “Tauopathies”[All Fields] OR “Alzheimer Disease”[All Fields] OR “Diffuse Neurofibrillary Tangles with Calcification”[All Fields] OR “Supranuclear Palsy, Progressive”[All Fields] OR “TDP-43 Proteinopathies”[All Fields] OR “Amyotrophic Lateral Sclerosis”[All Fields] OR “Frontotemporal Lobar Degeneration”[All Fields] OR “Frontotemporal Dementia”[All Fields] OR “Primary Progressive Nonfluent Aphasia”[All Fields]))
3. Search terms modified to include different variations of disease names: (QSM OR “quantitative susceptibility mapping”) AND ((“Chronic Traumatic Encephalopathy”[All Fields] OR “Heredodegenerative Disorder”[All Fields] OR Alexander*[All Fields] OR “Amyloid Neuropath*”[All Fields] OR “Bulbo-Spinal Atrophy”[All Fields] OR Canavan*[All Fields] OR Cockayne*[All Fields] OR “Dystonia Musculorum Deformans”[All Fields] OR “Gerstmann-Straussler-Scheinker Disease”[All Fields] OR “Hepatolenticular Degeneration”[All Fields] OR “Hereditary Central Nervous System Demyelinating Diseases”[All Fields] OR “Hereditary Sensory and Autonomic Neuropath*”[All Fields] OR Dysautonomia*[All Fields] OR “Hereditary Sensory and Motor Neuropathy”[All Fields] OR Alstrom*[All Fields] OR “Charcot-Marie-Tooth”[All Fields] OR “Giant Axonal Neuropathy”[All Fields] OR Refsum*[All Fields] OR “Spastic Paraplegia”[All Fields] OR Huntington*[All Fields] OR Lafora*[All Fields] OR “Myotonia Congenita”[All Fields] OR “Myotonic Dystrophy”[All Fields] OR Neurofibromato*[All Fields] OR “Neurofibromatosis 1”[All Fields] OR “Neurofibromatosis 2”[All Fields] OR “Neuronal Ceroid-Lipofuscinoses”[All Fields] OR “Hereditary Optic Atroph*”[All Fields] OR “Optic Atrophy”[All Fields] OR Leber*[All Fields] OR Wolfram*[All Fields] OR “Pantothenate Kinase-Associated Neurodegeneration”[All Fields] OR “Spinal Muscular Atroph*”[All Fields] OR “Spinocerebellar Degen*”[All Fields] OR Friedreich*[All Fields] OR “Myoclonic Cerebellar Dyssynergia”[All Fields] OR Olivopontocerebellar*[All Fields] OR Spinocerebellar*[All Fields] OR “Machado-Joseph Disease”[All Fields] OR Tourette*[All Fields] OR “Tuberous Sclerosis”[All Fields] OR “Unverricht-Lundborg”[All Fields] OR “Motor Neuron Disease”[All Fields] OR “Amyotrophic Lateral Sclerosis”[All Fields] OR “Bulbar Palsy”[All Fields] OR “Muscular Atrophy”[All Fields] OR “Bulbo-Spinal Atrophy”[All Fields] OR “Spinal Muscular Atroph*”[All Fields] OR “Nervous System Paraneoplastic Syndrome”[All Fields] OR “Anti-N-Methyl-D-Aspartate Receptor Encephalitis”[All Fields] OR “Limbic Encephalitis”[All Fields] OR “Myasthenia Gravis”[All Fields] OR “Lambert-Eaton”[All Fields] OR “Transverse Myelitis”[All Fields] OR “Opsoclonus-Myoclonus Syndrome”[All Fields] OR “Paraneoplastic Cerebellar Degeneration”[All Fields] OR “Paraneoplastic Polyneuropathy”[All Fields] OR “Postpoliomyelitis Syndrome”[All Fields] OR Prion[All Fields] OR “Bovine Spongiform Encephalopathy”[All Fields] OR “Gerstmann-Straussler-Scheinker”[All Fields] OR “Fatal Familial Insomnia”[All Fields] OR Kuru[All Fields] OR Scrapie[All Fields] OR “Chronic Wasting Disease”[All Fields] OR “Subacute Combined Degeneration”[All Fields] OR Synucleinopath*[All Fields] OR Lewy[All Fields] OR “Multiple System Atrophy”[All Fields] OR Parkinson*[All Fields] OR Tauopath*[All Fields] OR Alzheimer*[All Fields] OR “Diffuse Neurofibrillary Tangles with Calcification”[All Fields] OR “Progressive Supranuclear Palsy”[All Fields] OR “TDP-43”[All Fields] OR “Amyotrophic Lateral Sclerosis”[All Fields] OR “Frontotemporal Lobar Degeneration”[All Fields] OR “Frontotemporal Dementia”[All Fields] OR “Primary Progressive Nonfluent Aphasia”[All Fields] OR PKAN OR ALS OR MSA OR Frontotemporal* OR Eaton-Lambert))
4. (QSM OR “quantitative susceptibility mapping”) AND (neurodegen*) Same strategy is pursued in Embase, PsycInfo and Scopus, tailoring the search to map/explode to all neurodegenerative diseases in step 1. Search terms in steps 2, 3 and 4 will remain the same.

## Appendix 2. Data extraction form

**Appendix 2. Data extraction form:**
|  |  |  |
| --- | --- | --- |
| *Study title:* |  |  |
| *Year:* |  | *Author:* |
| *Neurodegenerative Disease* |  |  |
| *Sample size* |  |  |
| *Mean age* |  |  |
| *Gender* |  |  |
| *Disease characteristics* |  |  |
| *Field strength, model and manufacturer* |  | *MRI protocol* |
| *MRI sequences details* |  |  |
| *QSM processing* |  |  |
| *ROIs investigated and segmentation:* |  |  |
| *Findings:* |  |  |

## Appendix 3. Quality (risk of bias) assessment tool

Appendix 3. Quality (risk of bias) assessment tool:
| Criteria | Yes | No | Other<br>(CD, NR, NA)* |
| --- | --- | --- | --- |
| 1. Was the research question or objective in this paper clearly stated and appropriate? |  |  |  |
| 2. Was the study population clearly specified and defined? |  |  |  |
| 3. Did the authors include a sample size justification? |  |  |  |
| 4. Were controls selected or recruited from the same or similar population that gave rise to the cases (including the same timeframe)? |  |  |  |
| 5. Were the definitions, inclusion and exclusion criteria, algorithms or processes used to identify or select cases and controls valid, reliable, and implemented consistently across all study participants? |  |  |  |
| 6. Were the cases clearly defined and differentiated from controls? |  |  |  |
| 7. If less than 100 percent of eligible cases and/or controls were selected for the study, were the cases and/or controls randomly selected from those eligible? |  |  |  |
| 8. Was there use of concurrent controls? |  |  |  |
| 9. Were the investigators able to confirm the diagnosis of the investigated disease prior to the MRI scan? |  |  |  |
| 10. Were the QSM techniques defined, valid, reliable, and implemented consistently (including the same time period) across all study participants? |  |  |  |
| 11. Were the researchers responsible for segmentation, processing and analysis of QSM blinded to the case or control status of participants? |  |  |  |
| 12. Were key potential confounding variables measured and adjusted statistically in the analyses? If matching was used, did the investigators account for matching during study analysis? |  |  |  |

